# Transcriptomic profiling of cardiac tissues from SARS-CoV-2 patients identifies DNA damage

**DOI:** 10.1101/2022.03.24.22272732

**Authors:** Arutha Kulasinghe, Ning Liu, Chin Wee Tan, James Monkman, Jane E Sinclair, Dharmesh D Bhuva, David Godbolt, Liuliu Pan, Andy Nam, Habib Sadeghirad, Kei Sato, Gianluigi Li Bassi, Ken O’Byrne, Camila Hartmann, Anna Flavia Ribeiro dos Santo Miggiolaro, Gustavo Lenci Marques, Lidia Zytynski Moura, Derek Richard, Mark Adams, Lucia de Noronha, Cristina Pellegrino Baena, Jacky Y Suen, Rakesh Arora, Gabrielle T. Belz, Kirsty R Short, Melissa J Davis, Fernando Souza-FonsecaGuimaraes, John F Fraser

## Abstract

The severe acute respiratory syndrome coronavirus 2 (SARS-CoV-2) is known to present with pulmonary and extra-pulmonary organ complications. In comparison with the 2009 pandemic (pH1N1), SARS-CoV-2 infection is likely to lead to more severe disease, with multi-organ effects, including cardiovascular disease. SARS-CoV-2 has been associated with acute and long-term cardiovascular disease, but the molecular changes govern this remain unknown.

In this study, we investigated the landscape of cardiac tissues collected at rapid autopsy from SARS-CoV-2, pH1N1, and control patients using targeted spatial transcriptomics approaches. Although SARS-CoV-2 was not detected in cardiac tissue, host transcriptomics showed upregulation of genes associated with DNA damage and repair, heat shock, and M1-like macrophage infiltration in the cardiac tissues of COVID-19 patients. The DNA damage present in the SARS-CoV-2 patient samples, were further confirmed by γ−H2Ax immunohistochemistry. In comparison, pH1N1 showed upregulation of Interferon-stimulated genes (ISGs), in particular interferon and complement pathways, when compared with COVID-19 patients.

These data demonstrate the emergence of distinct transcriptomic profiles in cardiac tissues of SARS-CoV-2 and pH1N1 influenza infection supporting the need for a greater understanding of the effects on extra-pulmonary organs, including the cardiovascular system of COVID-19 patients, to delineate the immunopathobiology of SARS-CoV-2 infection, and long term impact on health.

## Introduction

Since 2019, SARS-CoV-2 has led to the recognition of the emergence of broad-spectrum multi-organ disease with increasing prevalence of cardiac injury in hospitalized patients. SARS-CoV-2 has been associated with a wide range of cardiac complications. Specifically, the initial infection stage has been associated with acute myocardial injury^1-3^ with abnormal echocardiography findings in both left and right ventricles^4^, arrhythmias^1,2^, palpitations^5^, myocarditis^6^, heart failure^7^ and other *de novo* problems^8,9^. Paralleling these clinical symptoms is a growing body of evidence of increased cardiac complications in patients who have recovered from COVID-19^10^. Signs of persistent cardiac-related issues among individuals who have suffered from severe disease during the acute infection appear to be relatively common. These include continued chest pain^11^, palpitations^12^, abnormal left ventricular function^13^, myocardial infarction^13^, late gadolinium enhancement (LGE)^13,14^, and ischaemia^13^. Ongoing myocardial inflammation and LGE^15^ have also been reported even in patients who recovered from relatively mild, or even asymptomatic, COVID-19. Individuals who have recovered from mild COVID-19 suffer an increased burden of arrhythmias, chest pain, heart failure and vascular complications compared to uninfected patients, and this sequelae has been accompanied by excess use of drug therapies^16^. Currently, it remains unclear how long-term cardiac complications from COVID-19 will persist in convalescent patients. Studies have shown that the risk and 1-year burden of cardiovascular disease is substantial in survivors of acute COVID-19 ^17^.

Virus-induced cardiac complications are not unique to SARS-CoV-2 infection. Influenza A virus (IAV) infection has been frequently associated with myocardial injury and infarction, endocarditis, tachycardia, ST segment echocardial changes and atrial fibrillation^18,19^, mostly resolving within a year of infection^19^. The mechanisms by which respiratory viruses may cause cardiac complications are manifold and are also not fully characterised^20^. Thus, it is currently unclear if SARS-CoV-2 and IAVs induce these complications via similar or distinct pathways.

Evidence of SARS-CoV-2 direct cardiac infection remains equivocal. *In vitro* studies show infection and replication within human pluripotent stem cell-derived (hPSC) cardiomyocytes^21,22^, while hPSC-smooth muscle cells remain uninfected^23^. Autopsies of 39 patients with COVID-19 detected SARS-CoV-2 negative sense RNA indicative of active viral replication in the myocardium of only 5 patients with the highest viral loads^24^. In autopsies of a further 41 patients, SARS-CoV-2^+^ cells in the myocardium were rare despite viral RNA being detected in 30 hearts^25^. Furthermore, smaller studies of autopsy specimens from five and eight COVID-19 patients failed to detect SARS-CoV-2 within the heart entirely, even when patients displayed severe myofibrillar anomalies^21,26^. In IAV infection, it is equally uncertain whether early cardiomyocyte damage is linked primarily to virus presence or a secondary consequence of the immune response in IAV infection. IAV has been reported to replicate within hPSC-cardiomyocytes^27^, and IAV or antigens have been found directly in the heart in mice^27,28^ and humans^29,30^ in small case studies. In contrast, a larger US study of patients with acute viral myocarditis identified IAV RNA in cardiac samples from only 5 of 624 patients (0.8%) using PCR analyses^31^. Therefore, the contribution of direct cardiac viral infection to SARS-CoV-2 and IAV-induced cardiac complications remains both contentious and unclear.

One potential mechanistic disparity between influenza virus and SARS-CoV-2 induction of cardiac complications is the induction of an interferon (IFN) response in the heart. For example, two days post-IAV infection, the expression of IFN stimulated genes within the heart of IAV-infected mice was increased 50-fold in infected compared with uninfected mice^32^. In contrast, several studies have found that both respiratory and extra-respiratory type I interferon responses are significantly blunted during SARS-CoV-2 infection^33,34^. For example, a Syrian hamster model of SARS-CoV-2 infection showed inhibited type I IFN responses in the respiratory tract despite a high burden of replicating virus, accompanied by inflammation in the heart, which also lacked type I IFN upregulation^35^. This may be attributable to SARS-CoV-2 production of proteins which suppress type I IFN release^36,37^. The lack of efficient IFN induction by SARS-CoV-2 may be responsible for triggering the observed higher rate of cardiac complications than in more IFN-stimulatory seasonal IAVs^38,39^. Indeed, several studies have shown that type I IFNs play a protective role in the development of cardiovascular diseases such as pathological hypertrophy and virally-induced left ventricular dysfunction^40,41^. However, this hypothesis remains to be confirmed.

Transcriptomic analysis of patient myocardial tissue offers a unique opportunity to understand the mechanisms of SARS-CoV-2 and IAV induced cardiac complications. Specifically, spatial transcriptomics that consider intra-organ heterogeneity^42,43^, provide a powerful tool for characterising host responses to respiratory viral infections outside of the respiratory tract. Here, we use targeted spatial transcriptomic characterisation of myocardial tissue to generate an in-depth picture of the myocardial transcriptional landscape of COVID-19, pandemic H1N1 influenza and uninfected control patients and shed light on the mechanisms that might drive these different clinical outcomes. Our study revealed that DNA damage pathways were enriched for in COVID-19 tissues, whereas pH1N1 elicited a more inflammatory response in cardiac tissues.

## Methods

### Study Design

Myocardial tissues were obtained from patients at the Pontificia Universidade Catolica do Parana PUCPR in accordance with the National Commission for Research Ethics (CONEP) under ethics approval numbers: protocol number 3.944.734/2020 (for COVID-19 group), and 2.550.445/2018 (for pH1N1 and Control group). Families permitted the post-mortem biopsy of COVID-19 and H1N1pdm09 samples and conventional autopsy for the cases of the Control group. All SARS-CoV-2 and pH1N1 patients were confirmed for infection by RTqPCR of nasopharyngeal swab specimens. The study was ratified by The University of Queensland (UQ) Human Research Ethics Committee (HREC) (clearance number: 2020001792 / 30188020.7.1001.0020).

### Tissue preparation and histopathology

Tissue microarray (TMA) was constructed of single cores from 7 SARS-CoV-2, 2 pH1N1 and 6 control/healthy volunteer patients (Table 1) and cut onto positively charged slides (Bond Apex). Sections were stained by Fred Hutch pathology for hematoxylin and eosin (H&E) and Mason’s trichrome. Brightfield images were obtained using the Aperio (Leica Biosystems, US) slide scanner for histological assessment by a pathologist at The Prince Charles Hospital Pathology Laboratory.

**Table 1.**
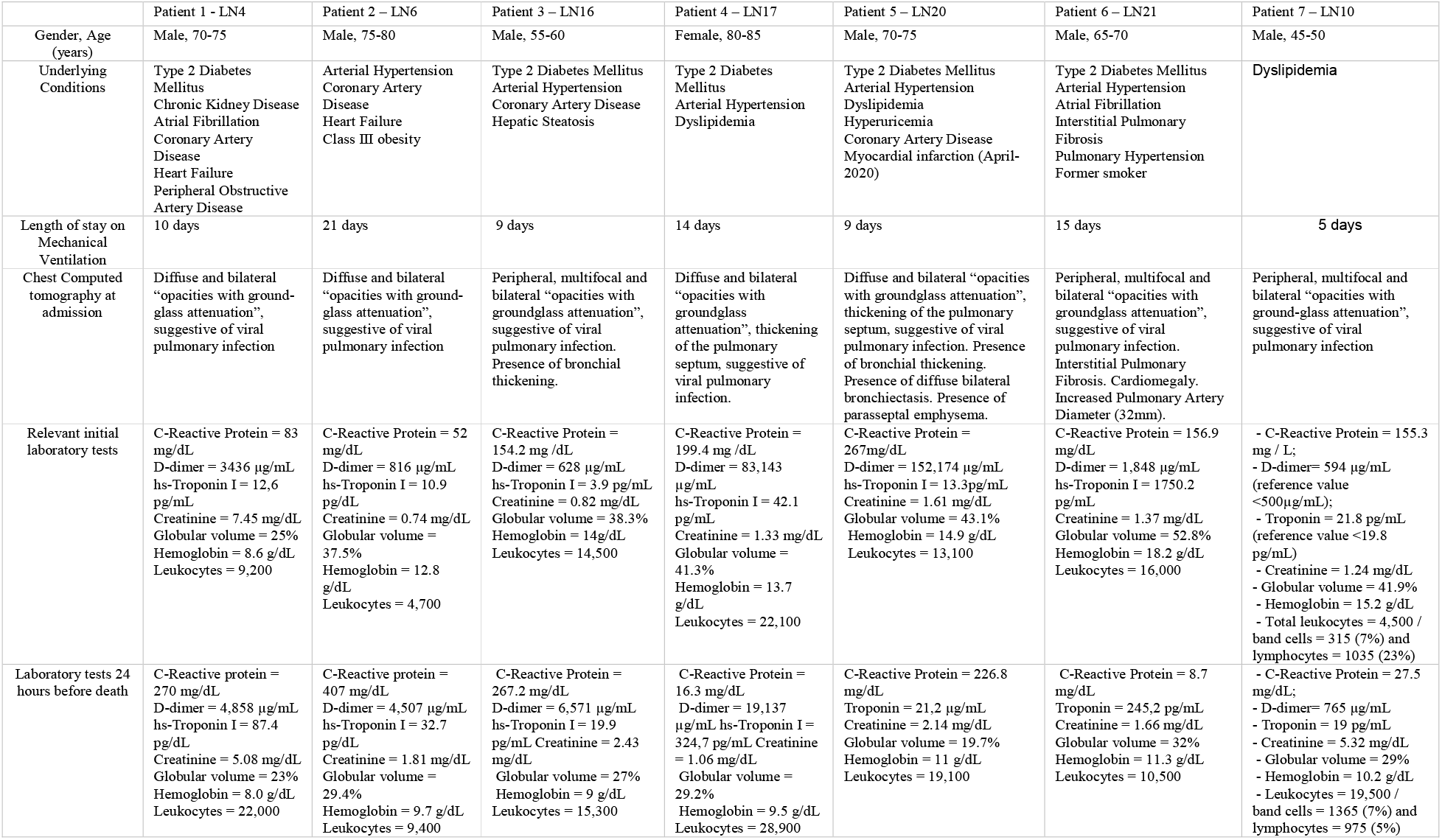
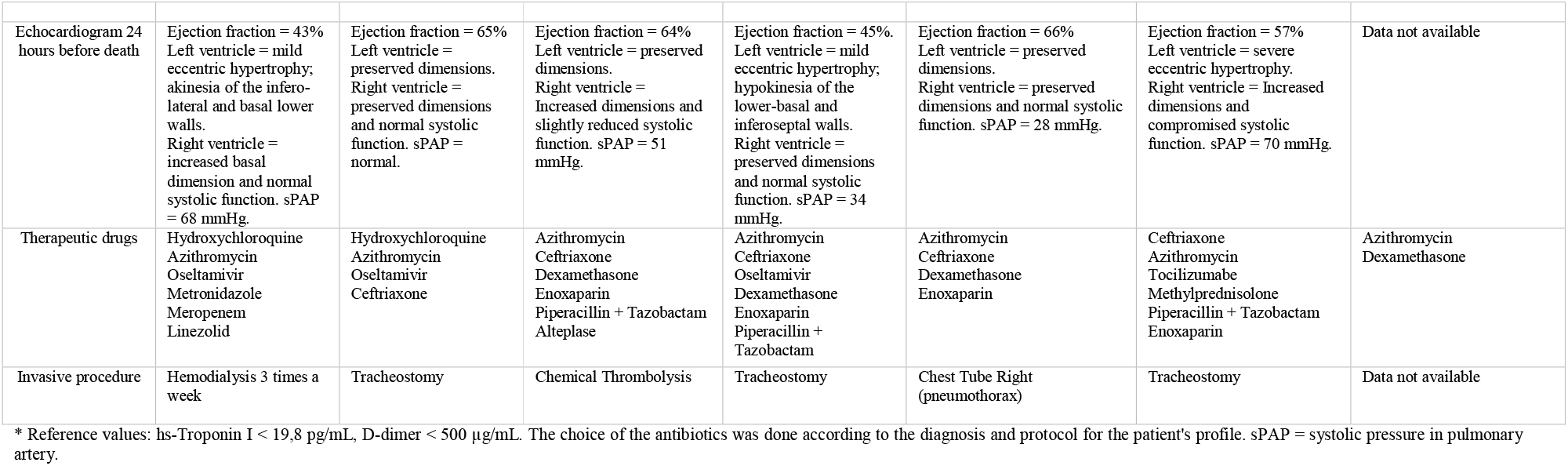
Clinicopathological findings for the COVID-19 patients

|  | Patient 1 - LN4 | Patient 2 – LN6 | Patient 3 – LN16 | Patient 4 – LN17 | Patient 5 – LN20 | Patient 6 – LN21 | Patient 7 – LN10 |
| --- | --- | --- | --- | --- | --- | --- | --- |
| Gender, Age (years) | Male, 70-75 | Male, 75-80 | Male, 55-60 | Female, 80-85 | Male, 70-75 | Male, 65-70 | Male, 45-50 |
| Underlying Conditions | Type 2 Diabetes Mellitus<br>Chronic Kidney Disease<br>Atrial Fibrillation<br>Coronary Artery Disease<br>Heart Failure<br>Peripheral Obstructive Artery Disease | Arterial Hypertension<br>Coronary Artery Disease<br>Heart Failure<br>Class III obesity | Type 2 Diabetes Mellitus<br>Arterial Hypertension<br>Coronary Artery Disease<br>Hepatic Steatosis | Type 2 Diabetes Mellitus<br>Arterial Hypertension<br>Dyslipidemia | Type 2 Diabetes Mellitus<br>Arterial Hypertension<br>Dyslipidemia<br>Hyperuricemia<br>Coronary Artery Disease<br>Myocardial infarction (April-2020) | Type 2 Diabetes Mellitus<br>Arterial Hypertension<br>Atrial Fibrillation<br>Interstitial Pulmonary Fibrosis<br>Pulmonary Hypertension<br>Former smoker | Dyslipidemia |
| Length of stay on Mechanical Ventilation | 10 days | 21 days | 9 days | 14 days | 9 days | 15 days | 5 days |
| Chest Computed tomography at admission | Diffuse and bilateral “opacities with ground-glass attenuation”, suggestive of viral pulmonary infection | Diffuse and bilateral “opacities with ground-glass attenuation”, suggestive of viral pulmonary infection | Peripheral, multifocal and bilateral “opacities with groundglass attenuation”, suggestive of viral pulmonary infection. Presence of bronchial thickening. | Diffuse and bilateral “opacities with groundglass attenuation”, thickening of the pulmonary septum, suggestive of viral pulmonary infection. | Diffuse and bilateral “opacities with groundglass attenuation”, thickening of the pulmonary septum, suggestive of viral pulmonary infection. Presence of bronchial thickening. Presence of diffuse bilateral bronchiectasis. Presence of paraseptal emphysema. | Peripheral, multifocal and bilateral “opacities with groundglass attenuation”, suggestive of viral pulmonary infection. Interstitial Pulmonary Fibrosis. Cardiomegaly. Increased Pulmonary Artery Diameter (32mm). | Peripheral, multifocal and bilateral “opacities with ground-glass attenuation”, suggestive of viral pulmonary infection |
| Relevant initial laboratory tests | C-Reactive Protein = 83 mg/dL<br>D-dimer = 3436 µg/mL<br>hs-Troponin I = 12.6 pg/mL<br>Creatinine = 7.45 mg/dL<br>Globular volume = 25%<br>Hemoglobin = 8.6 g/dL<br>Leukocytes = 9,200 | C-Reactive Protein = 52 mg/dL<br>D-dimer = 816 µg/mL<br>hs-Troponin I = 10.9 pg/dL<br>Creatinine = 0.74 mg/dL<br>Globular volume = 37.5%<br>Hemoglobin = 12.8 g/dL<br>Leukocytes = 4,700 | C-Reactive Protein = 154.2 mg /dL<br>D-dimer = 628 µg/mL<br>hs-Troponin I = 3.9 pg/mL<br>Creatinine = 0.82 mg/dL<br>Globular volume = 38.3%<br>Hemoglobin = 14g/dL<br>Leukocytes = 14,500 | C-Reactive Protein = 199.4 mg /dL<br>D-dimer = 83,143 µg/mL<br>hs-Troponin I = 42.1 pg/mL<br>Creatinine = 1.33 mg/dL<br>Globular volume = 41.3%<br>Hemoglobin = 13.7 g/dL<br>Leukocytes = 22,100 | C-Reactive Protein = 267mg/dL<br>D-dimer = 152,174 µg/mL<br>hs-Troponin I = 13.3pg/mL<br>Creatinine = 1.61 mg/dL<br>Globular volume = 43.1%<br>Hemoglobin = 14.9 g/dL<br>Leukocytes = 13,100 | C-Reactive Protein = 156.9 mg/dL<br>D-dimer = 1,848 µg/mL<br>hs-Troponin I = 1750.2 pg/mL<br>Creatinine = 1.37 mg/dL<br>Globular volume = 52.8%<br>Hemoglobin = 18.2 g/dL<br>Leukocytes = 16,000 | - C-Reactive Protein = 155.3 mg / L;<br>- D-dimer= 594 µg/mL (reference value <500µg/mL);<br>- Troponin = 21.8 pg/mL (reference value <19.8 pg/mL)<br>- Creatinine = 1.24 mg/dL<br>- Globular volume = 41.9%<br>- Hemoglobin = 15.2 g/dL<br>- Total leukocytes = 4,500 / band cells = 315 (7%) and lymphocytes = 1035 (23%) |
| Laboratory tests 24 hours before death | C-Reactive protein = 270 mg/dL<br>D-dimer = 4,858 µg/mL<br>hs-Troponin I = 87.4 pg/dL<br>Creatinine = 5.08 mg/dL<br>Globular volume = 23%<br>Hemoglobin = 8.0 g/dL<br>Leukocytes = 22,000 | C-Reactive protein = 407 mg/dL<br>D-dimer = 4,507 µg/mL<br>hs-Troponin I = 32.7 pg/dL<br>Creatinine = 1.81 mg/dL<br>Globular volume = 29.4%<br>Hemoglobin = 9.7 g/dL<br>Leukocytes = 9,400 | C-Reactive Protein = 267.2 mg/dL<br>D-dimer = 6,571 µg/mL<br>hs-Troponin I = 19.9 pg/mL.Creatinine = 2.43 mg/dL<br>Globular volume = 27%<br>Hemoglobin = 9 g/dL<br>Leukocytes = 15,300 | C-Reactive Protein = 16.3 mg/dL<br>D-dimer = 19,137 µg/mL<br>hs-Troponin I = 324.7 pg/mL<br>Creatinine = 1.06 mg/dL<br>Globular volume = 29.2%<br>Hemoglobin = 9.5 g/dL<br>Leukocytes = 28,900 | C-Reactive Protein = 226.8 mg/dL<br>Troponin = 21.2 µg/mL<br>Creatinine = 2.14 mg/dL<br>Globular volume = 19.7%<br>Hemoglobin = 11 g/dL<br>Leukocytes = 19,100 | C-Reactive Protein = 8.7 mg/dL<br>Troponin = 245.2 pg/mL<br>Creatinine = 1.66 mg/dL<br>Globular volume = 32%<br>Hemoglobin = 11.3 g/dL<br>Leukocytes = 10,500 | - C-Reactive Protein = 27.5 mg/dL;<br>- D-dimer= 765 µg/mL<br>- Troponin = 19 pg/mL<br>- Creatinine = 5.32 mg/dL<br>- Globular volume = 29%<br>- Hemoglobin = 10.2 g/dL<br>- Leukocytes = 19,500 / band cells = 1365 (7%) and lymphocytes = 975 (5%) |
| Echocardiogram 24 hours before death | Ejection fraction = 43%<br>Left ventricle = mild eccentric hypertrophy; akinesia of the infero-lateral and basal lower walls.<br>Right ventricle = increased basal dimension and normal systolic function. sPAP = 68 mmHg. | Ejection fraction = 65%<br>Left ventricle = preserved dimensions.<br>Right ventricle = preserved dimensions and normal systolic function. sPAP = normal. | Ejection fraction = 64%<br>Left ventricle = preserved dimensions.<br>Right ventricle = Increased dimensions and slightly reduced systolic function. sPAP = 51 mmHg. | Ejection fraction = 45%.<br>Left ventricle = mild eccentric hypertrophy; hypokinesia of the lower-basal and inferoseptal walls.<br>Right ventricle = preserved dimensions and normal systolic function. sPAP = 34 mmHg. | Ejection fraction = 66%<br>Left ventricle = preserved dimensions.<br>Right ventricle = preserved dimensions and normal systolic function. sPAP = 28 mmHg. | Ejection fraction = 57%<br>Left ventricle = severe eccentric hypertrophy.<br>Right ventricle = Increased dimensions and compromised systolic function. sPAP = 70 mmHg. | Data not available |
| Therapeutic drugs | Hydroxychloroquine<br>Azithromycin<br>Oseltamivir<br>Metronidazole<br>Meropenem<br>Linezolid | Hydroxychloroquine<br>Azithromycin<br>Oseltamivir<br>Ceftriaxone | Azithromycin<br>Ceftriaxone<br>Dexamethasone<br>Enoxaparin<br>Piperacillin + Tazobactam<br>Alteplase | Azithromycin<br>Ceftriaxone<br>Oseltamivir<br>Dexamethasone<br>Enoxaparin<br>Piperacillin + Tazobactam | Azithromycin<br>Ceftriaxone<br>Dexamethasone<br>Enoxaparin | Ceftriaxone<br>Azithromycin<br>Tocilizumabe<br>Methylprednisolone<br>Piperacillin + Tazobactam<br>Enoxaparin | Azithromycin<br>Dexamethasone |
| Invasive procedure | Hemodialysis 3 times a week | Tracheostomy | Chemical Thrombolysis | Tracheostomy | Chest Tube Right (pneumothorax) | Tracheostomy | Data not available |

\* Reference values: hs-Troponin I < 19,8 pg/mL, D-dimer < 500 µg/mL. The choice of the antibiotics was done according to the diagnosis and protocol for the patient's profile. sPAP = systolic pressure in pulmonary artery.

### Immunohistochemistry and RNAscope (SARS-CoV-2)

Immunohistochemistry was performed on a Leica Bond-RX autostainer (Leica Biosystems, US) with antibody targeting SARS-CoV-2 spike protein (Abcam, ab272504) at 2 μg·mL^−1^. Heat induced epitope retrieval was performed in buffer ER1 at 100°C for 20 min, and signal visualised with 3,3⍰-Diaminobenzidine (DAB) substrate. Slides were imaged using a Zeiss Axioscanner (Carl Zeiss, Germany). RNAscope® probes (ACDbio, US) targeting SARS-CoV-2 spike mRNA (nCoV2019, #848561-C3), ACE2 host receptor mRNA (#848151-C2), and host serine protease TMPRSS2 mRNA (#470341-C1) were used as per manufacturer instructions for automation on Leica Bond RX. DNA was visualised with Syto13 (Thermofisher Scientific), channel 1 with Opal 570 (1:500), channel 2 with Opal 620 (1:1500), and channel 3 with Opal 690 (1:1500) (PerkinElmer). Fluorescent images were acquired with Nanostring Mars prototype DSP at 20x. Anti-gamma H2A.X staining was performed as above on autostainer with antibody at 4ug/ml (ab26350). Antigen retrieval was performed in buffer ER1 at 100°C for 10 min, and signal was visualised with Opal 520. Slides were scanned on Vectra Polaris (Akoya Biosciences, US) and unmixed in Inform (Akoya Biosciences, US). Image analysis was performed in QuPath^44^ to generate H-scores for γ-H2AX nuclear staining.

### Nanostring Digital Spatial Profiling (DSP): COVID-19 immune atlas panel

A serial section TMA slide, was freshly sectioned and prepared according to the Nanostring GeoMX Digital Spatial Profiler (DSP) slide preparation for RNA profiling (Nanostring, US). Briefly, slides were baked 1 h at 60°C and then processed by Leica Bond RX autostainer. Slides were pre-treated with Proteinase K and then hybridised with mRNA probes in the COVID-19 Immune Atlas panel with additional SARS-CoV-2 probe panel. After incubation, slides were washed and then stained with αSMA, CD3, CD68, and Syto83 for 1 h then loaded into the Nanostring GeoMX DSP instrument for scanning and ROI selection. ROI selection was guided by morphology markers to capture similar tissue structures across tissue cores where possible. Oligonucleotides linked to hybridised mRNA targets were cleaved and collected for counting using Illumina i5 and i7 dual indexing. PCR reactions were performed with 4 μl of a GeoMx DSP sample. AMPure XP beads (Beckman Coulter) were used at 1.2× bead-to-sample ratio for PCR product purification. Paired-end sequencing (2×75) was performed using NextSeq550 up to 400M total aligned reads. Fastq files were processed by DND system and uploaded to GeoMX DSP system where raw and Q3 normalised counts of all targets were aligned with regions of interest (ROIs).

### Transcriptomic data analysis

Data used in this study result from a mRNA assay conducted with the NanoString’s GeoMx COVID-19 Immune atlas panel using the GeoMX Digital Spatial Profiler (DSP). The data were measurements of RNA abundance of over 1800 genes, including 22 add-in COVID-19 related genes, 4 SARS-CoV-2 specific genes and 2 negative control (SARS-CoV-2 Neg, NegProbe) genes and 32 internal reference genes. Transcriptomic measurements were made on regions of interests within each core. Control samples are COVID-19-free and pH1N1-free. In total, 48 ROIs were analysed (16 COVID-19, 4 pH1N1 and 28 Control). Factors considered in this dataset include disease type (COVID-19, pH1N1 and Control), patient of origin, dominant tissue type (Blood vessel, mixed vessel/myocardium, myocardium only).

### Bioinformatics analyses

#### Data exploration and quality control

Data exploration and quality controls were conducted on negative probe-QC count data generated from the Covid-19 Immune atlas DSP mRNA assay. Data were first rescaled into log2-transformed count per million (CPM) data to account for library size variation, followed by computing relative log expression (RLE) and principal components analysis (PCA) of the scaled count data to assess the overall distribution, factor variance from the experimental design and the presence of unwanted batch effects across all datasets.

#### Batch correction and normalization

The dataset comprise of two different experiments, leading to effects of batch between experiments that is required to be considered and eliminated. Batch correction and normalisation requires negative control genes which were derived from RNA-seq count data of atrial appendage, left ventricle and aorta tissues from the Genotype-Tissue Expression (GTEx) project^45^ (2017-06-05_v8_RNASeQCv1.1.9). Coefficient of variance (CV) was calculated for each gene after the transformation of raw count to log-scaled CPM count to account for library size variation. Genes were then sorted based on a z-score transformation. The top 500 heart related GTEx stable genes were intersected with the CTA panel genes used in this study, resulting in 32 overlapped genes (Supplementary Table 1). The list of negative control genes was further curated with 7 genes of potential biological relevance and 3 genes with a high CV (mean CV (log transformed) over 3.5) within each batch were removed from the list (**Supplementary Table 1 and Supplementary Figure 1**), leaving 22 genes as negative control genes for downstream analyses. *RUV4* from the ruv R package^46^ was then used to for the normalisation using the 22 negative control genes and the factor k set to 3. The result of the normalisation was then assessed by computing RLE and PCA of the normalised count data.

#### Differential expression analysis

Differential expression (DE) analysis was performed using R packages edgeR^47^ (v3.34.0) and limma^48^ (3.48.0). Briefly, differential expression was modelled using linear models with experimental factors as predictors. The variation in gene expression was modelled as the combination of a common dispersion that applies to all genes and a gene-specific dispersion. To estimate the common and gene-wise variation, the variation of each gene was modelled by borrowing information from all other genes using an empirical Bayes approach while treating the variation of patients as a random effect using *DuplicateCorrelation* in Limma. The linear model was then fitted to a given experimental design containing the biological factors of interest and the weight matrix from *RUV4* normalisation to account for unwanted variations. DE was performed for distinct contrasts of interest. The resulting statistic was an empirical Bayes moderated t-statistic which was more robust than a t-statistic from a classic t-test. In this study the main factor of interest is disease with the main contrasts investigated as COVID19 vs control, pH1N1 vs control and COVID19 vs pH1N1

#### Gene set enrichment analysis

Gene sets from the Molecular Signatures Database^49^ (MsigDB, v7.2) Hallmarks, C2 (curated gene sets) and C5 (gene ontology terms) categories and KEGG pathways gene sets were obtained using the *getMisgdb* and *appendKEGG* functions from the msigdb R package (v1.1.5)^49^. Gene set enrichment analysis (GSEA) were performed using *fry* from the limma package. False discovery rate (FDR) of 0.05 was set as the threshold to determine significantly enriched gene sets. Taking advantage of the fact that most gene sets are related and share genes in common, the results of the GSEA were interrogated and visualised by an unbiased approach using a novel network enrichment and visualization R package vissE^50^. The vissE approach exploits this geneset-geneset overlap relationship to enable interpretation of clusters of gene sets and further summarize results. This approach allows natural clustering of the gene sets to take place, so that significant perturbations can be better visualized while allowing the other less obvious, sensible but un-expected results to be highlighted. To inspect the concordance between the transcriptomic profile of each ROI and DNA damage-related gene set, we used R package singscore^51^.

## Results

Cardiac tissue was obtained from 7 SARS-CoV-2 infected patients, 2 pH1N1 influenza patients and 6 control patients at rapid autopsy. Patients were confirmed as having SARS-CoV-2 infection by RTqPCR of nasopharyngeal swabs and confirmed ‘glass opacities’ characteristic of pulmonary infection. The SARS-CoV-2 cohort was composed of 6 male and 1 female with a mean age of 69 years (range 46-81) (Table 1). These patients had a number of comorbidities, including type II diabetes, cardiac disease and arterial hypertension. All patients underwent mechanical ventilation ranging from 5 to 21 days. Tissue immunohistochemistry showed oedema in all SARS-CoV-2 infected patients and myocarditis in one patient. Pathology review of the myocardial histology samples were indifferent to patients of similar age and co-morbidities. All COVID-19 samples were found to have RNAscope SARS-CoV-2 near background levels and considered not to have viral RNA present.

### Spatial transcriptomic analysis

Nanostring DSP analysis was performed on (n=7) SARS-CoV-2, (n=2) pH1N1 and (n=6) control patients across 2 tissue microarrays. Region of interest (ROI) selection was performed to enable the capture of targeted transcriptome from sufficient myocardial tissue (>200 cells) to generate robust count data. Regions selected included blood vessels (endothelial cells, aSMA+), myocardial muscle (striated tissue) and mixed regions containing both vessels and surrounding muscle tissue (**Figure 2**). Bar codes targeting these regions were collected to generate morphology marker informed compartments. The barcodes were sequenced, mapped, and counted by NGS readout as per manufacturer’s instructions. Quality control QC was performed to remove outlying probes and collapse counts from 5 genes per probe to single gene measurements. The QC files were output for further bioinformatics analysis.

**Figure 1.**
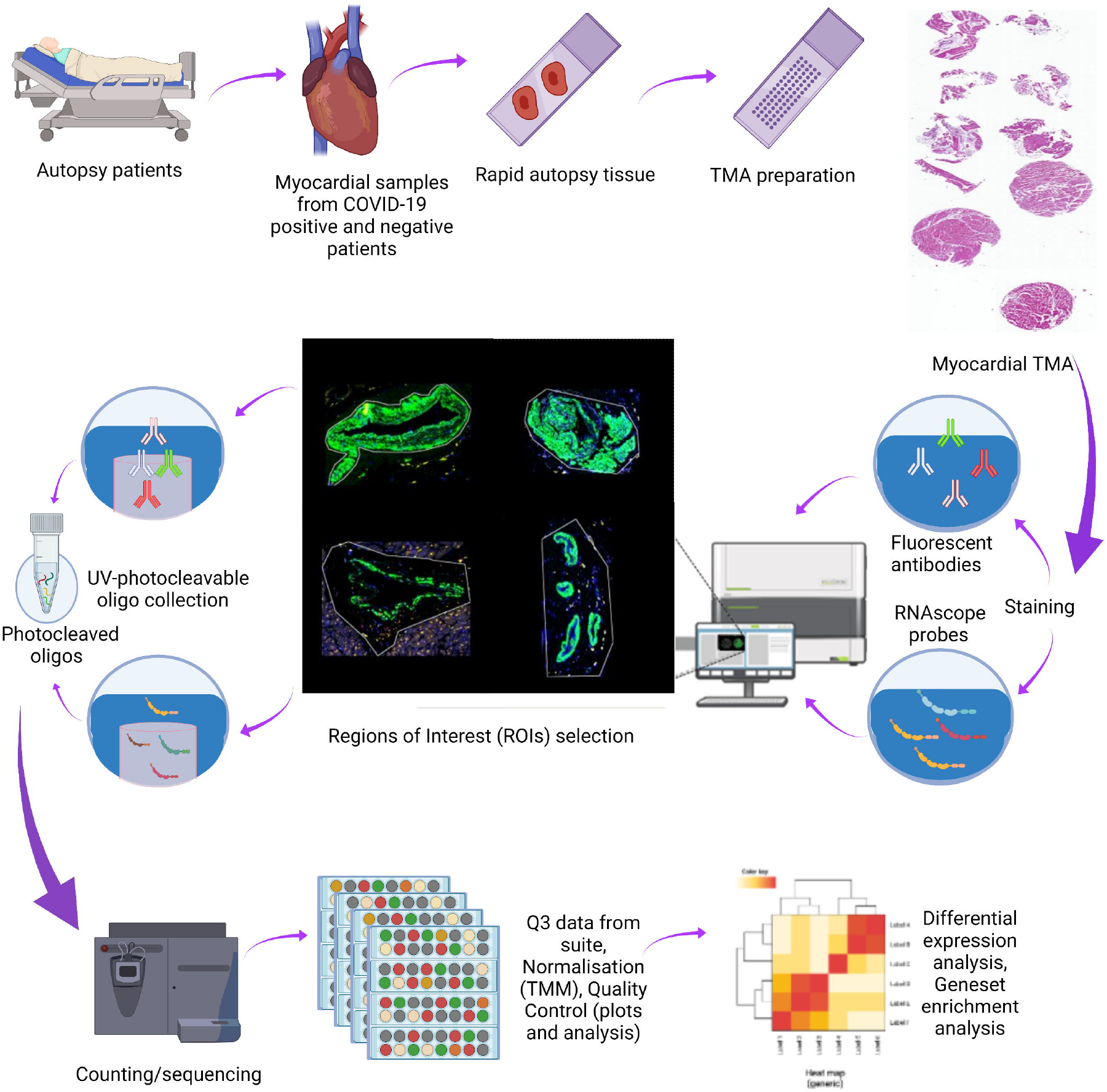
Study Schema. Cardiac tissues were collected from COVID-19 and pH1N1 patients at rapid autopsy. Samples were prepared onto tissue microarrays and profiled using targeted spatial transcriptomics (Immune Atlas Panel, Nanostring Technologies). Myocardial, blood vessels and mixed populations were captured using ‘region of interest’ selection strategies to liberate the transcript data. These data were counted by next generation sequencing to obtain digital expression counts per region of interest.

**Figure 2.**
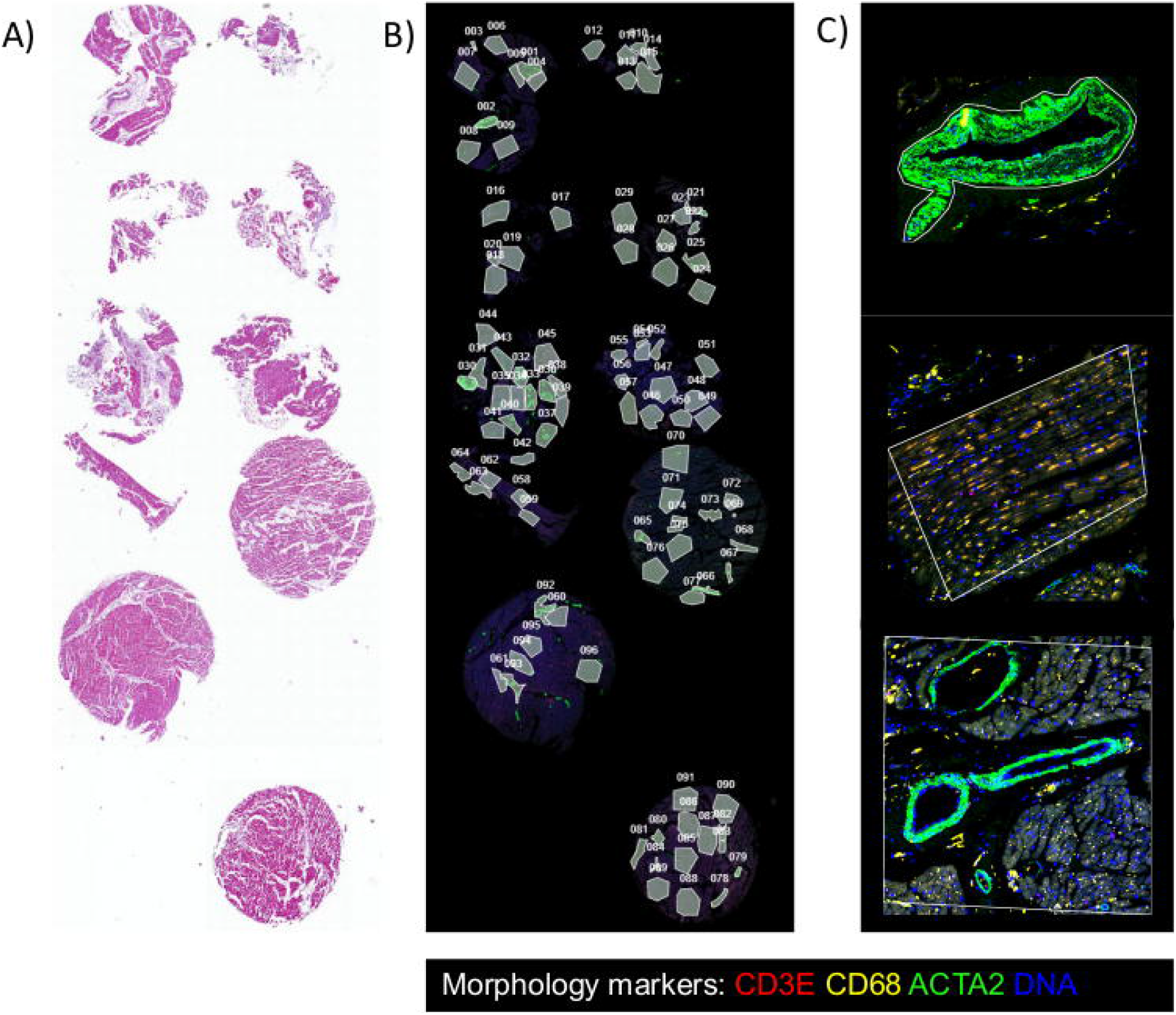
Representative Immunohistochemistry. (A) H&E staining of the tissue microarray (B) Regions of interest selected for spatial profiling by the Nanostring GeoMX Digital Spatial Profiler (DSP) assay (C) Regions of interest for the blood vessel, myocardium, and mixed vessel/myocardium. Morphology markers for CD3E, CD68 and ACTA2 shown here.

### Data Standardisation and Batch Correction

The batch correction adjusted for effects from the two independent batches of tissue microarray data. This can be seen from the principal component analysis (PCA) (**Figure 3A and 3B**) where control samples from two batches are not clustered until the batch correction was performed. Additional to batch effect, technical variations of the data were mostly removed after batch correction, which can be visualised in the relative log expression (RLE) plots (**Figure 3D and 3E**), which are known to be sensitive to technical variation while insensitive to biological variation (REF).

**Figure 3.**
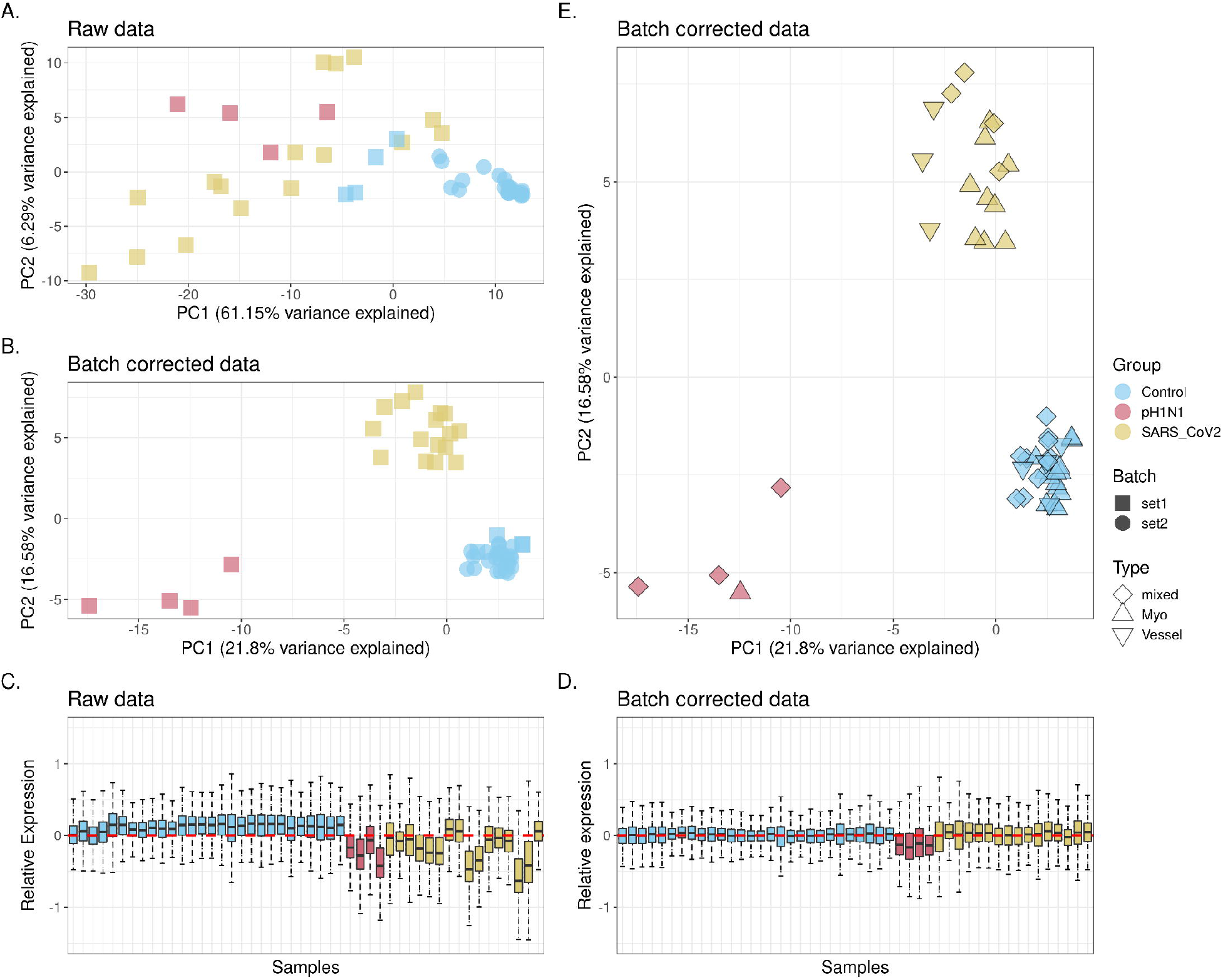
Batch correction and variability assessment of the spatial transcriptomic data. Principal component (PCs) analysis identifies the variability from batch effect in the transcriptomic data before (A) and after (B) batch correction. Relative log expression (RLE) plots (D and E) show the removal of technical variations after batch correction. Variabilities contributed by biological factors are visualised (C) in PC1 and PC2 of the batch corrected data.

In terms of biological variance (**Figure 3B**), differences in disease types are mainly explained by the variance from PC1 and PC2, while consistent local clustering of different tissue types can be observed within each disease type cluster. However, slight patient effect can still be observed, especially in control samples (**Supplementary Figure 2**), which is required to be account for in the downstream analysis.

### Differential expression analysis

To determine the virus induced effects on the cardiac tissues, we performed differential expression analysis of (A) COVID-19 vs pH1N1, (B) COVID-19 vs control and (C) pH1N1 vs control (**Figure 4 and Supplementary Table 2**). Notably, there is an increased expression of interferon response gene expression in pH1N1 patients compared with COVID-19 patients (**Figure 4A**). This is shown by an increase in interferon-induced transmembrane proteins (*IFITM1, IFITM2, IFIT3*) and interferon stimulating genes (ISGs). In contrast, in COVID-19, increased expression of chemokine ligands such as *CCL15* (also known as macrophage inflammatory protein 5) were found, which has a chemotactic effect eliciting a transient increase of intracellular calcium are contributes towards inflow of monocytes/macrophages and neutrophils ^52^. Moreover, *CCL15* and *SSX1* (a member of the synovial sarcoma X breakpoint protein family) were also significantly differentially expressed in COVID-19 vs control tissues (**Figure 4B**). *SSX1* has been previously found to be present in lung tissues with high amounts of SARS-CoV-2 RNA ^53^, and thought to be induced by SARS-CoV-2 as an adaptive response to interferon. *HSPA1A*, of the family of heat shock proteins, HSP70*s*, a classical molecular chaperone, was found to be upregulated in COVID-19 vs control, and is thought to elicit a potent anti-inflammatory effect ^54^. In pH1N1 vs control (Figure 4C), a strong anti-viral, interferon response was observed.

**Figure 4.**
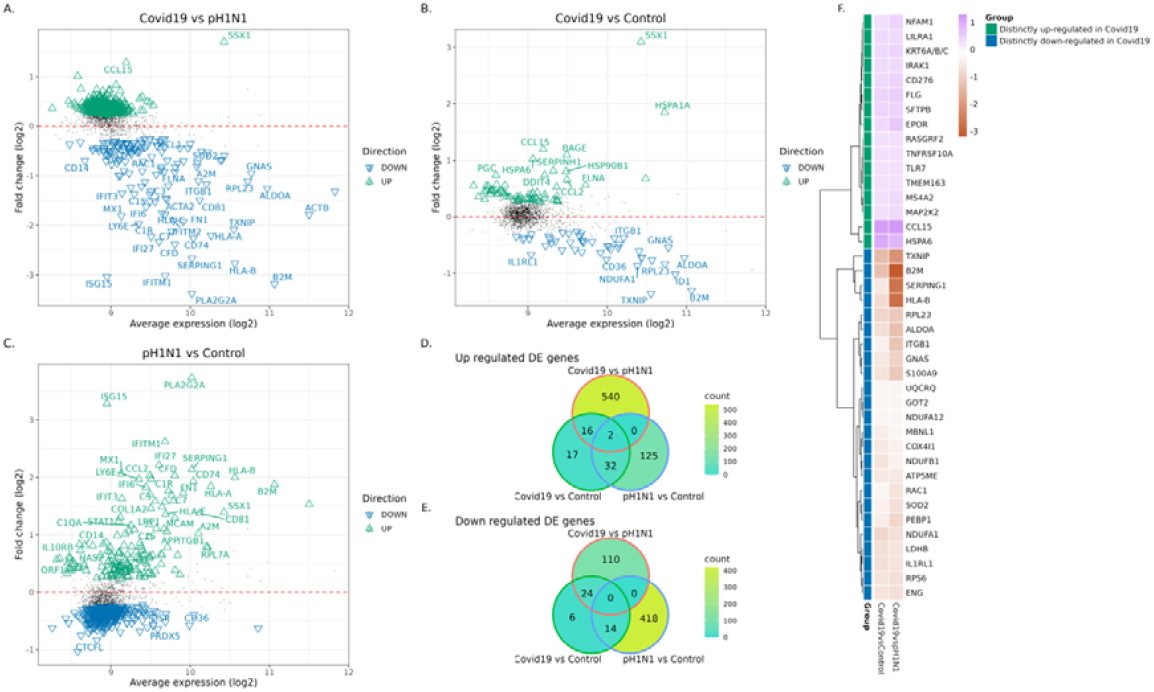
Differential expression analysis. Distribution of differentially expressed (DE) genes as a function of the average transcript expression (log2) and fold change (log2) identified in the following comparisons were visualised: (A) COVID-19 samples versus pH1N1 samples (B) COVID-19 samples versus Control samples, and (C) pH1N1 samples versus Control. Green triangles indicate up-regulated, blue triangles down-regulated, and black dots indicate non-DE genes. Differential expression genes were derived using voom-limma pipeline with limma: *duplication Correlations* and FDR threshold with Benjamini Hochberg adjusted p-value < 0.05. Venn diagram (D and E) is used to visualise the intersection of DE genes from each comparison. Heatmap (F) is used to show the fold change (log2) of the DE genes that are distinctly up- or down-regulated in Covid19 samples.

Comparison of the overlapping DE genes between these comparisons (see Venn diagrams, in **Figure 4D and E**) suggested 16 and 24 unique DE genes are up- and down-regulated, respectively, in COVID-19 tissues compared with both pH1N1 and control samples. These 40 genes were not found to be DE between pH1N1 vs control samples, suggesting these as (**Figure 4F**) distinctly regulated for COVID-19 patients. Of these COVID-19 specific genes, the inflammatory response-related gene NFAM1 and TNF receptor gene *TNFRSF10A* are found to be up-regulated in concordance with previous reports suggesting up-regulation in the throat swab samples ^55^ and in T cells ^56^ of COVID-19 patients, respectively. Of the downregulated genes, of particular interest is *IL1RL1* (which encodes the ST2 protein and is involved in the IL-33/ST2 signalling pathway with its receptor cytokine IL-33. This gene has been known to be cardioprotective for myocardial functions and has been implicated with increased cardiomyocyte hypertrophy and ventricular fibrosis upon germline deletion of ST2 in mouse models. Furthermore, ST2^57^ has also been tolled as a promising prognostic biomarker for COVID-19^58^ with ST2 serum level suggested to be significantly increased in COVID-19 patients ^59^, as opposed to the significant downregulation of *IL1RL1 in COVID-19 heart samples*.

### Gene set enrichment analysis

We performed gene set enrichment analysis (GSEA) on differentially expressed (DE) genes of each patient groupings (COVID19, pH1N1 and control) and visualised gene set enrichment by computing a similarity network from lists of curated gene sets using vissE^50^ (**Figure 5**). For the comparison of COVID-19 against pH1N1 samples, the gene set similarity network was visualised as a network (**Figure 5B**). We then identified higher-order phenotypic changes associated with interferon responses in the clustering of gene sets (**Figure 5A**). Specifically for interferon responses, cluster 15 (**Figure 5A&B**, blue boundary) represents gene sets such as *reactome interferon alpha beta signalling, interferon response* and *interferon induced antiviral module* that are down-regulated in SARS-CoV-2 samples relative to pH1N1 samples (Supplementary Table 3). Within this cluster, interferon-related genes including *IFI27, IFIT3* and *IFITM1* (**Figure 5C**, cluster 14) were all found to be downregulated.

**Figure 5.**
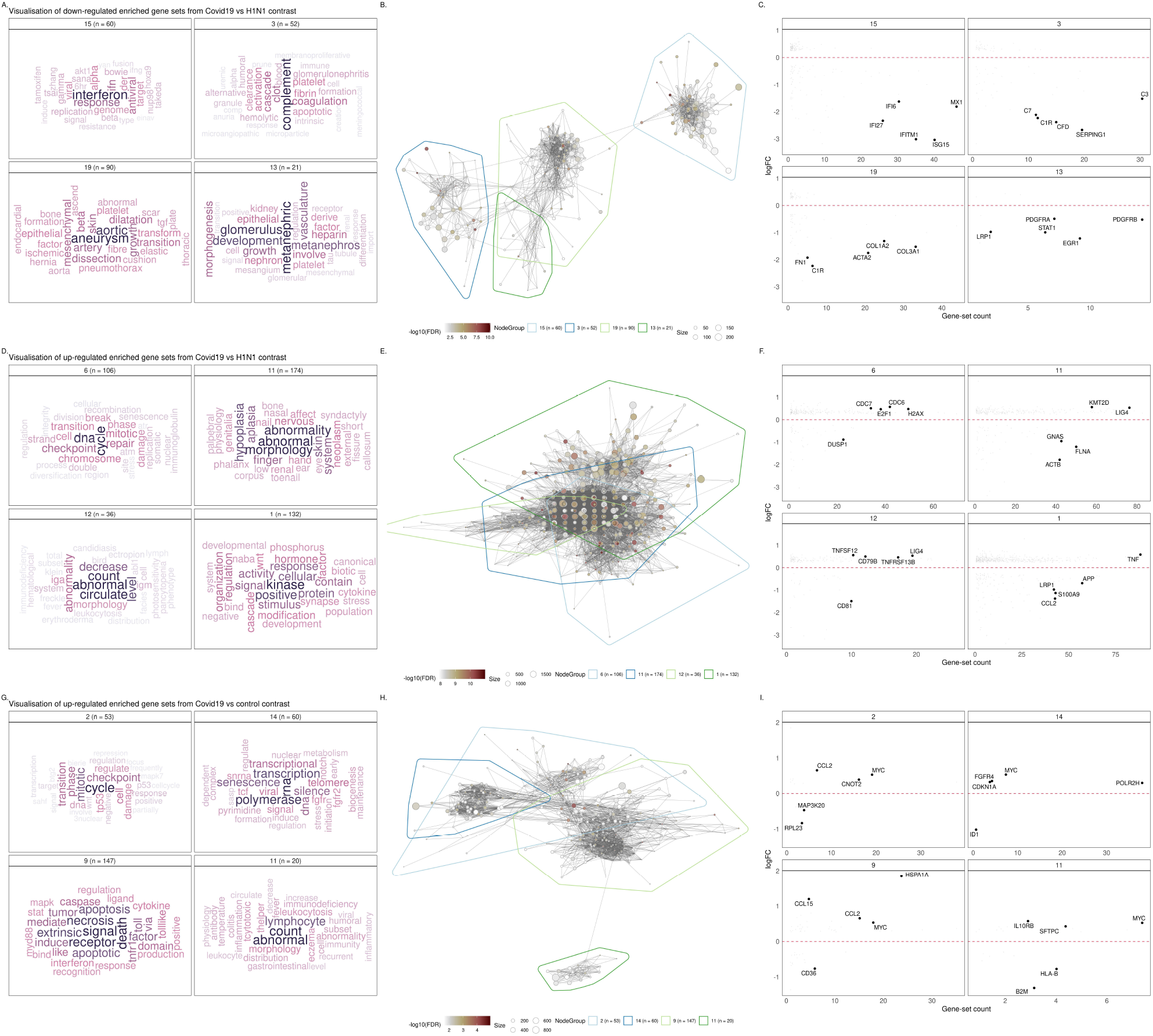
Visualisation of significantly enriched gene sets different comparisons. A, D and G: Cluster annotations based on text-mining analysis of gene set names. 9 gene-set clusters representing biological themes of each comparison are displayed. B, E and H: Gene-set overlap graphs of gene sets enriched in up/down-regulated DE genes in different comparisons with nodes representing gene sets and edges representing overlaps based on the Jaccard Index. Nodes are coloured based on the significance of enrichment. C, F and I: Fold-change (log2-scaled) for genes belonging to gene sets in the cluster plot against the number of gene-sets in the cluster the gene belongs to.

Visualising downregulated gene sets comparing COVID-19 to pH1N1, identified complement activation pathway was downregulated in Cluster 3 (**Figure 5A-C**) for COVID-19, reflected by initiator complement factor C1 complex, enzymatic mediator C3, membrane attach complex C7, amongst other genes in this pathway (**Figure 5A-C**, cluster 3). *SERPING1* which encodes the C1 inhibitor (C1I) which inhibits C1r and C1s of the first complement component, is strongly downregulated in COVID-19 compared with pH1N1 infection. Mutations of C1 are associated with dysregulation of the complement pathway and angioedema. The complement system is made up of about 40 heat-stable plasma proteins which are involved in seven functional components, and cross-talk in a catalytic cascade. Activation of the complement pathway is critical for immune homeostasis and deficiency can lead to life-threatening infections ^60^.

The most predominant gene sets upregulated in COVID-19 when compared with pH1N1 (**Figure 5, D-F**), were DNA break, damage and repair, cellular abnormality, and cell cycle, including checkpoint and signalling (**Figure 5, D-F**). This trend in cellular damage was seen in the COVID-19 compared with normal cardiac tissue and showed enrichment of cell death and senescence gene sets (**Figure 5, G-I**). In addition, leukocyte and myeloid cell gene sets were also enriched for in COVID-19 patients compared with pH1N1 and control cardiac tissues, reflecting the higher number of neutrophils and leukocytes observed in severe COVID-19 patients.

To further investigate and visualise the GSEA outcome of our transcriptomic data, staining for γ-H2AX was performed ^61^, an established marker of DNA damage. Two of five COVID-19 tissues exhibited significant nuclear γ-H2AX signal, indicating the detection of DNA damage in these tissues. The degree of positive staining across these samples also coincided with the enrichment of GSEA scores of DSP regions for 6 DNA damage gene sets (Supplementary Figure 3).

## Discussion

Pandemic H1N1 influenza drives a cytokine storm of generalised inflammation disrupting the heart which presents with fever, tachycardia, and arrhythmias. In contrast, COVID-19 drives a now recognised syndrome which can result in acute myocardial infarction, myocardial injury, heart failure, disseminated thrombosis, hypotension, arrhythmias and sudden cardiac death ^17^.

In COVID-19 patients with adverse outcomes, cardiac troponin I and brain-type natriuretic peptide (BNP) are elevated in ICU admission patients^62^. However, while biomarker changes are indicative of tissue damage, the mechanisms involved in cardiac injury have not been fully established. Recent studies have shown that myocarditis is prevalent in COVID-19, however, evidence for subclinical cardiac inflammation or mechanisms regulating this process has been limited.

In influenza, viral binding to host cells induces a type I and III interferon response, including inflammatory cytokines (IL-6, TNF-α) and chemokines. The binding of the type I and III IFNs to their receptors results in activation of the JAK/STAT pathways for the induction of ISGs such as IFI27. This can lead to acute myocarditis, a common complication of influenza infection^63^. In contrast to influenza infection, induction of these pathways in COVID-19 is low^33^, a feature supported by recent autopsy studies in which virus was not detected as a cause of myocarditis^64^. It was previously observed that excessive signalling induced by type 1 IFN induces inflammation-driven myocardial infarction and this can be triggered by self-DNA release and activation of the cGAS-STING-IRF3 pathway. This pathway is also connected to inflammation-induced DNA damage, which was suggested by others and by our current dataset^65^.

DNA damage response and repair mechanisms have been involved in the pathogenesis of chronic conditions such as diabetes and cardiovascular disease^66^. However, the role of SARS-CoV-2 in inducing genome instability has not been fully ascertained. *In vitro* studies have shown that the SARS-CoV-2 spike protein inhibits DNA damage repair by impeding the recruitment of DNA damage repair checkpoint proteins, a pre-requisite for V(D)J recombination in adaptive immunity. This has further been confirmed in spike protein over expressed cells by upregulation of DNA damage marker γ-H2AX 67.

Key clusters of genes impacted were uniquely altered by SARS-CoV-2 infection and were distinct from pH1N1. These focus on DNA damage and repair pathways and the consequent cell cycle arrest pathways. Notably we observed upregulation of LIG4, an ATP-dependent DNA ligase which acts to repair DNA double-strand breaks via the non-homologous enjoining pathway^68^. LIG4 expression is known to be enhanced following DNA damage and by Wnt/β-catenin signalling^69^, suggesting that COVID-19-induced DNA damage might be responsible for induction of LIG4 in cardiac tissue. While this remains to be determined, the helicase NSP13 protein expressed by the related SARS-CoV-1 is known to induce DNA damage and replication fork stress by interacting directly with DNA polymerase *δ*^70^. Given the NSP13 protein shares 99.8% sequence homology between SARS-CoV and SARS-CoV-2, it is possible that infection may induce DNA damage within myocardial tissue. However, SARS-CoV-2 infection has been observed, at least *in vitro*, to induce telomere shortening^71^. This feature is attributed with senescence which aligns with the upregulation of this gene set pathway in COVID-19 myocardial tissues in our study. Interestingly, telomere stability is controlled by the DNA damage response proteins, as a telomere resembles a DNA break. Shortened telomeres result in a persistent DNA damage response, although at this point the function of these foci are unknown ^72^.

In cardiac tissues, we also observed that COVID-19 induced downregulation of gene clusters involved in in mitochondrial function and metabolic regulation. Mitochondrial dysfunction is linked with COVID-19 whereby SARS-CoV-2 viral proteins interact with host mitochondrial proteins^73^. For example, viral open reading frame 9c (ORF9c) interacts with NDUFAF1 and NDUFAB1^73,74^, genes we identified in our study that are required for cellular bioenergetics as part of Complex I. Indeed, SARS-CoV-2 manipulation of mitochondrial activity is likely to enable evasion of mitochondrial-mediated innate immunity^74,75^.

Dysfunctional mitochondria are also associated with myocarditis^76^, and persistent inflammation causing irreversible myocardium damage^77,78^. Damage to the myocardium is triggered by danger-associated molecular patterns (DAMPs) which are recognised by toll-like receptors (TLR) that are expressed on immune and heart parenchymal cells^78,79^. Consistently, we observe upregulation of gene clusters associated with TLR signalling in heart tissue. Crucially, mitochondrial lipid, peptides and circulating mitochondrial DNA (mtDNA) are a source of DAMPs^76^. For example, increased circulating mtDNA is detected following myocardial infarction^80^ and can cause TLR-induced cardiomyocyte death^81^. Pertinently, antibodies against a key mitochondrial lipid, cardiolipin, have also been reported following serological testing of a critically ill COVID-19 patient exhibiting thrombocytopenia and coagulopathy^82^. Indeed, the pathways and gene sets identified in our study point to a key role for SARS-CoV-2-induced cardiac injury. However, further work is warranted to discern whether direct SARS-CoV-2 infection of cardiac tissue or other physiological events are responsible for the cardiac injury observed in our cohort.

Our study provides a comprehensive complex cellular blueprint across the full composition of cardiac tissues responding to SARS-CoV2 and H1N1 influenza using highly sophisticated spatio-temporal analyses. This study is limited by the number of samples for each cohort, in particular for the pH1N1 group. In addition, this analysis was restricted to autopsy samples which are unlikely to reflect the full spectrum of COVID-19 disease.

More comprehensive assessments of post-acute sequelae are needed to determine the short and long-term impacts of SARS-CoV-2 infection. It is known that DNA damage and impaired repair mechanisms foster genome instability and are involved in several chronic diseases. Long-term studies are needed to identify new onset heart disease from the early, and even subclinical, lesions as time post infection transpires.

## Data Availability

All data produced in the present study are available upon reasonable request to the authors

## Acknowledgements

The authors thank Miki Haraguchi and Stephanie Weaver (Fred Hutchinson Cancer Research Pathology) and WEHI (previously known as the Walter and Eliza Hall Institute) Histology core for their assistance. This research was carried out at the Translational Research Institute, which is supported by a grant from the Australian Government.

---

**Supplementary Figure 1.**
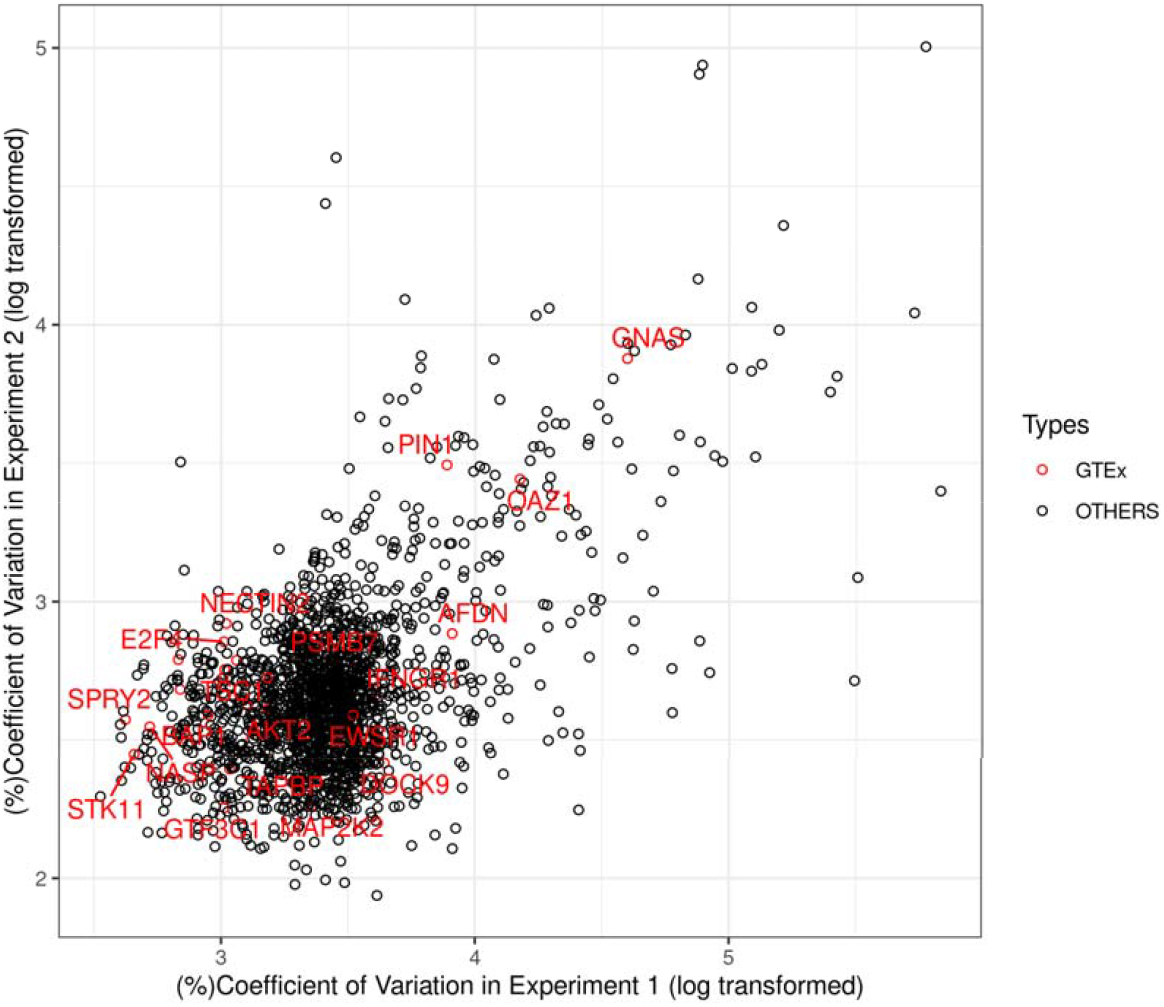
The identification of negative control genes from the dataset. Coefficient of variance (log-transformed) of all genes within each batch are computed, genes that are identified as GTEx stable genes are highlighted.

**Supplementary Figure 2.**
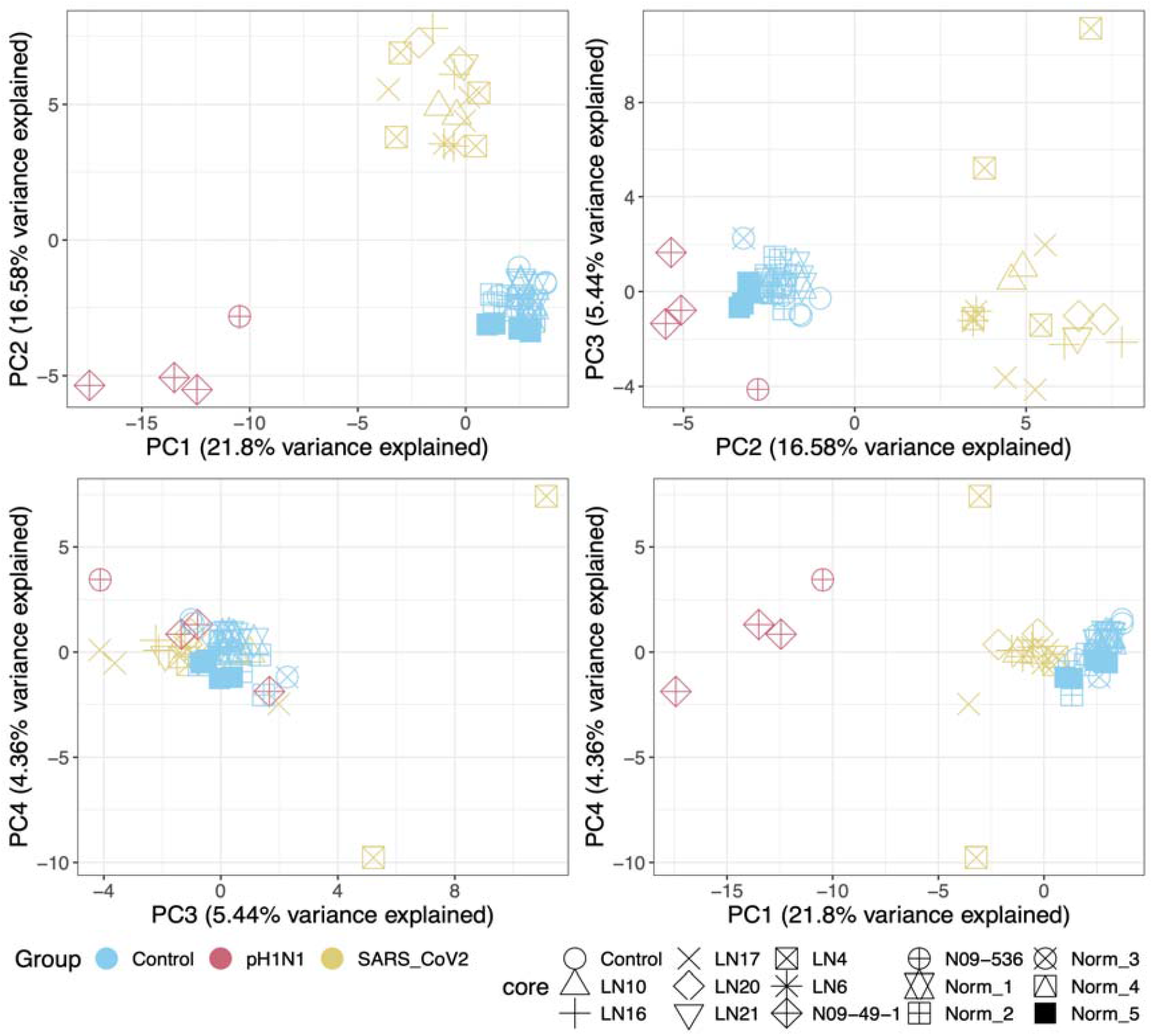
Pair-wise PCA plot of batch-corrected data.

**Supplementary Figure 3.**
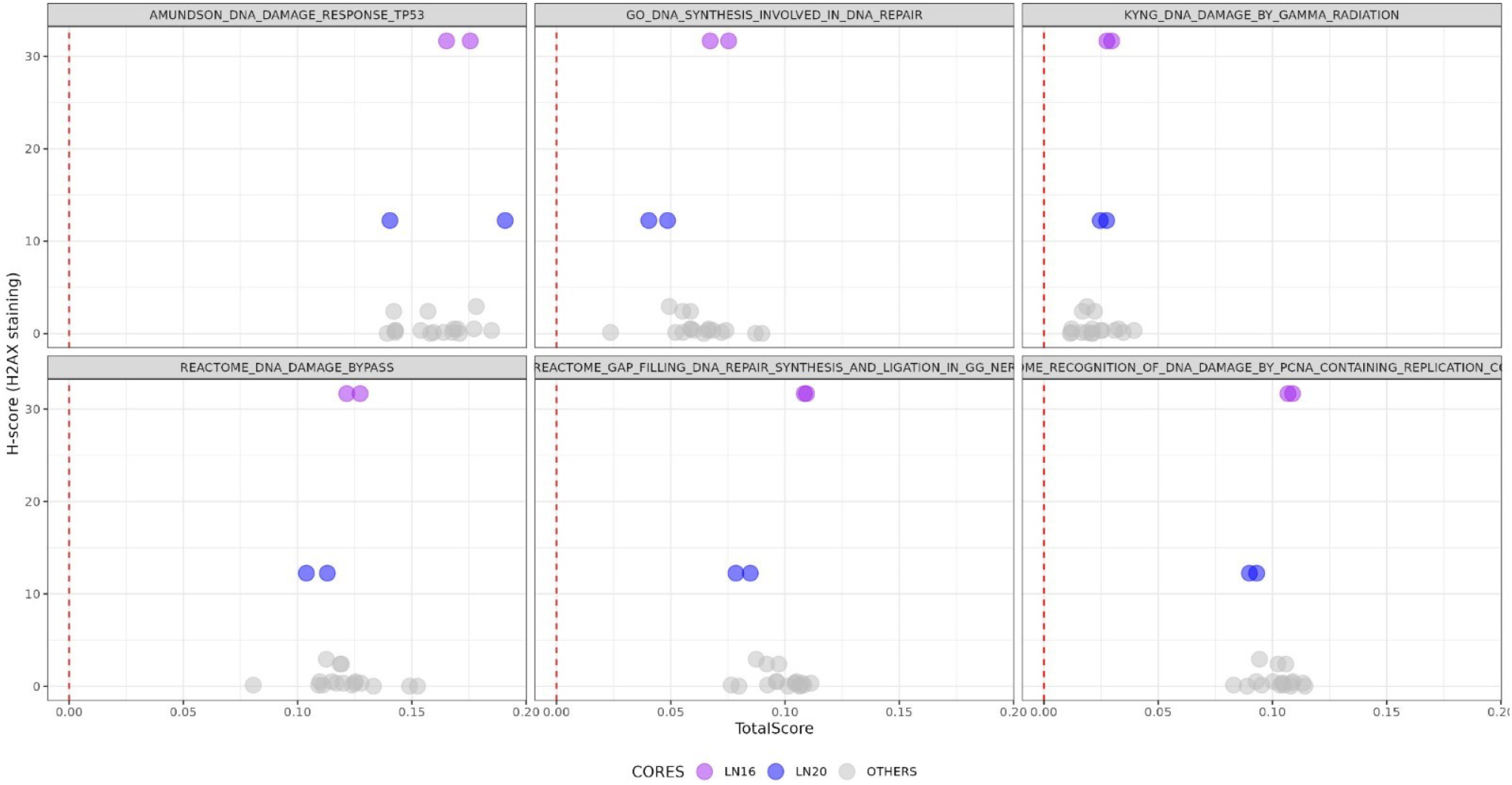
Gamma H2AX staining corresponds with GSEA scores of DSP regions. Two COVID-19 cores, LN16, LN20 (coloured), exhibited positive staining. DSP regions within these cores were scored for enrichment of DNA damage GSEA gene sets using singscore. High levels of gamma-H2AX in LN20 corresponded with a positive shift in score in GO DNA synthesis involved and in DNA damage/repair pathways.

**Supplementary Table 1**. GTEx heart-specific stable genes and the negative control genes.

**Supplementary Table 2**. Differential expression analysis.

**Supplementary Table 3**. Gene set enrichment analysis.

