## Supplementary figures and images for "Transcriptomic profiling of cardiac tissues from SARS-CoV-2 patients identifies DNA damage"

### SupplementaryFigure1 (1).png

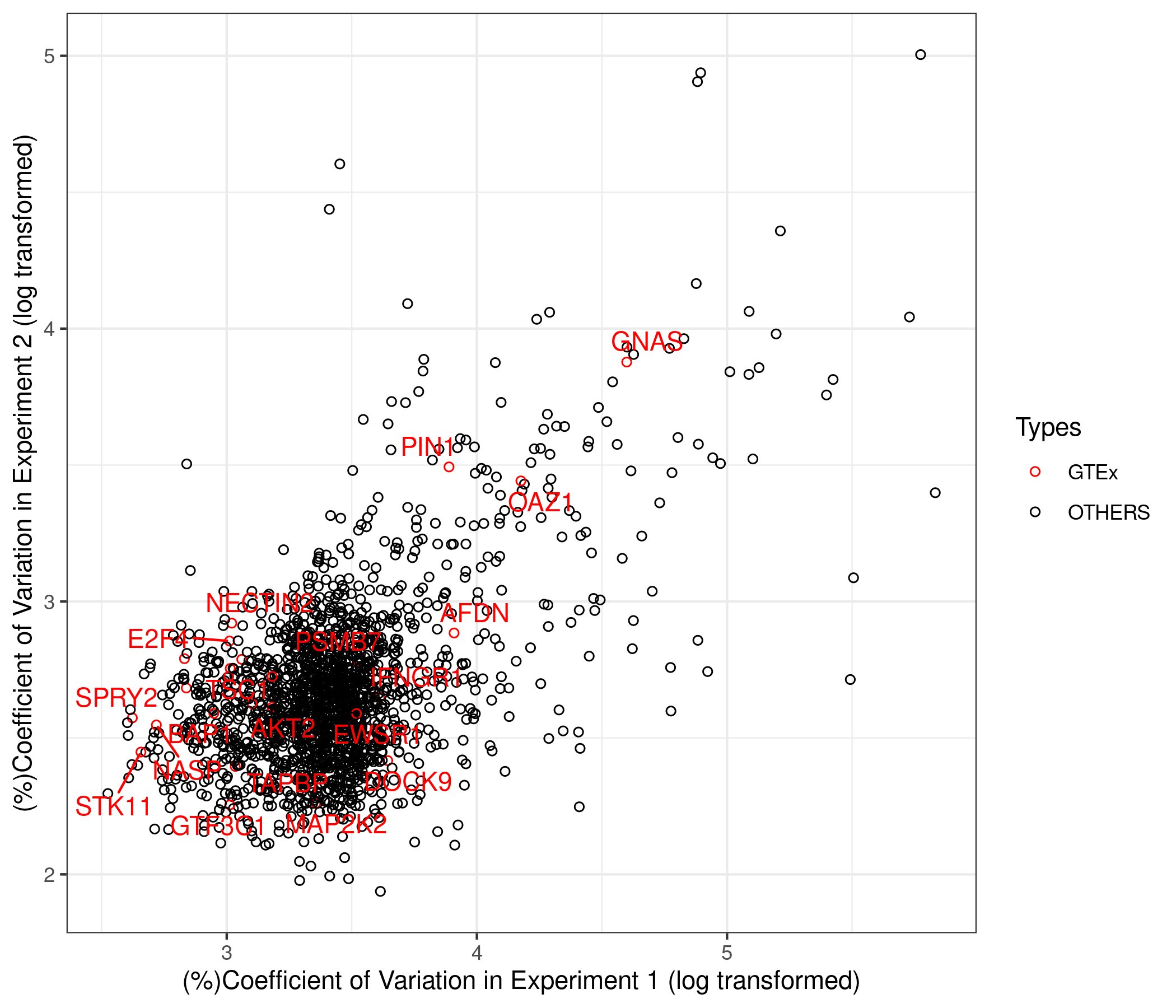

### SupplementaryFigure2.jpg

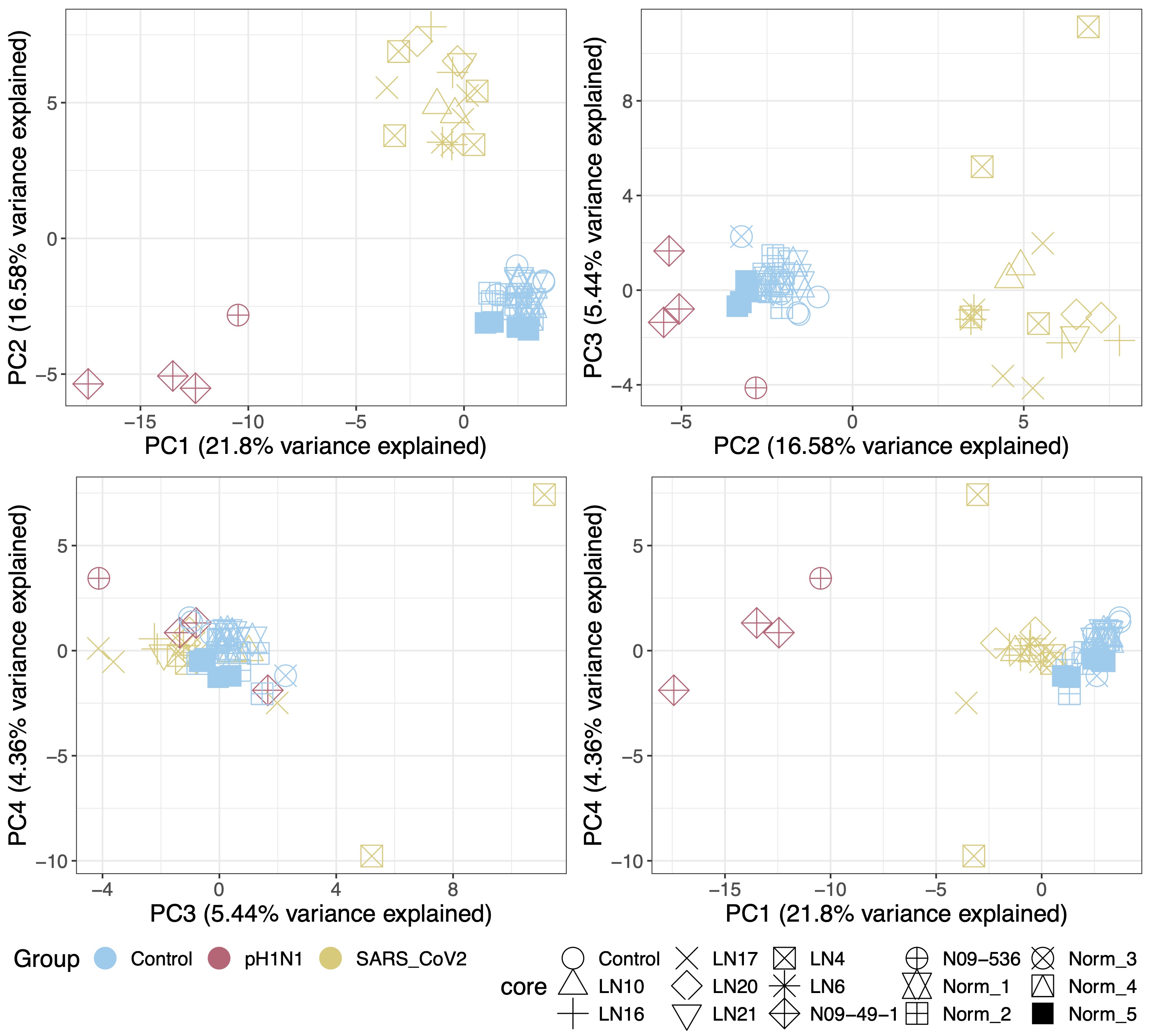

### SupplementaryFigure3.jpg

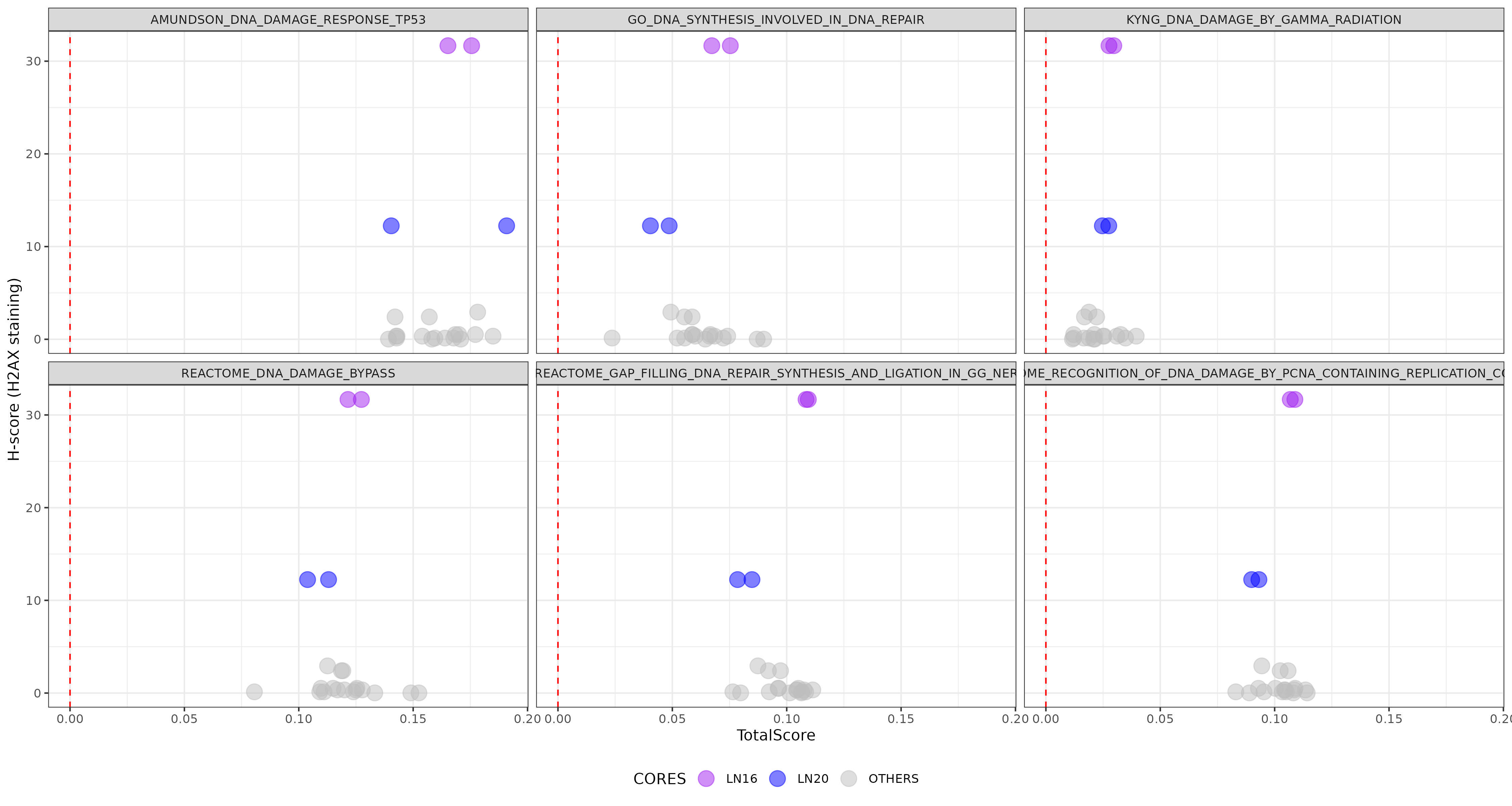
